# Household costs and catastrophic health expenditure from malaria in pregnancy in Western Kenya

**DOI:** 10.64898/2026.09.21.26363610

**Authors:** Amani Thomas Mori, Silke Fernandes, Kara Hanson, Hellen C. Barsosio, Frederick Omiti, Alloys K’Oloo, Michael A. Ojuok, Dawn Odiwa, Benson Omondi, Elizabeth Okello, Miriam Taegtmeyer, Simon Kariuki, Mwayi Madanitsa, John Lusingu, Franklin Mosha, Duolao Wang, Daniel Minja, Victor Mwapasa, Kamija S. Phiri, Kenneth Maleta, Jayne Webster, Jenny Hill, Feiko O. ter Kuile, Bjarne Robberstad

## Abstract

**Background:** Malaria in pregnancy (MiP) imposes a substantial economic burden on households in sub-Saharan Africa. We estimated household costs of MiP and their distribution across wealth quintiles among HIV-uninfected pregnant women in western Kenya receiving intermittent preventive treatment (IPTp) with sulfadoxine-pyrimethamine (SP), dihydroartemisinin-piperaquine (DP), or dihydroartemisinin-piperaquine plus provider education.

**Methods:** This study was nested within a three-armed, pragmatic, open-label, cluster-randomised trial across 15 health facilities over 10 months. Exit interviews were conducted during the second and third trimesters, approximately 5–6 months after the start of the intervention period. Direct and indirect household costs from the most recent malaria episode were compared with income to estimate catastrophic health expenditure (CHE). Costs were analysed using mixed-effects generalised linear model and their distribution across wealth quintiles using the concentration index. Cost data collected in 2020 (Kenyan shillings) were inflated and converted to 2024 US dollars.

**Findings:** Among 1,189 participants, 247 (21%) self-reported having sought care for MiP (212 outpatients, 24 inpatients, 11 unspecified). Mean costs were USD 24.9 for outpatient and USD 57.1 for inpatient care per episode. Indirect costs accounted for 87% and 70% of outpatient and inpatient costs, respectively. The mean outpatient malaria costs were equivalent to six or more days while inpatient costs were half a month or more of the minimum wages in the study area. The incidence of CHE for outpatient care was 8%, and neither outpatient costs nor CHE varied between wealth quintiles (concentration index =0.02, p=0.571 and 0.01, p=0.304, respectively).

**Conclusions:** MiP imposes a substantial economic burden on households in western Kenya, despite a national policy of free maternal health services. Most costs arose from productivity losses rather than payments for care; financial protection therefore depends on the prevention of malaria episodes.

## Introduction

Malaria in pregnancy (MiP) affects 13 million pregnancies annually in sub-Saharan Africa, causing severe maternal and neonatal complications [1, 2]. Intermittent preventive treatment in pregnancy (IPTp) with sulfadoxine-pyrimethamine (SP) is recommended by WHO for prevention of MiP; however, *Plasmodium falciparum* resistance to SP has reduced its effectiveness in East and southern Africa [3-5]. Monthly IPTp with the long-acting dihydroartemisinin-piperaquine (IPTp-DP) is more efficacious [6] and more cost-effective than IPTp-SP in these settings [7]. However, IPTp-SP was shown to be more effective than IPTp-DP in reducing adverse pregnancy outcomes, possibly related to non-antimalarial effects of SP on these outcomes [6].

Malaria also imposes severe social and economic burdens on households and health systems and has been associated with catastrophic health expenditure and poverty [8, 9]. Household costs may include transportation, direct medical expenses and indirect costs from lost productivity. The economic burden of malaria has been estimated at 3-8% of the household budget in affected countries [10]. A recent study across four countries (Democratic Republic of Congo, Madagascar, Mozambique and Nigeria) estimated that the cost of outpatient episodes of MiP ranged from USD 17 to USD 31, while inpatient costs ranged from USD 36 to USD 61. Indirect costs accounted for over two-thirds of the total costs [11].

The high cost of care-seeking are well-known barriers to maternal and child health interventions in low-and middle-income countries [12, 13] and contribute to inequities in IPTp-SP coverage [14, 15]. Poor, uneducated, and rural women have the lowest IPTp coverage [16, 17] and disproportionately bear the health and economic burden of MiP. Therefore, this study aimed to estimate the household costs of MiP among HIV-uninfected pregnant women receiving monthly IPTp with SP, DP, and DP plus provider education. The study also examined whether household cost burdens varied by socioeconomic status. The comparative cost-effectiveness of the IPTp strategies is reported separately.

## Methods

This study was conducted as part of a three-armed, pragmatic, open-label, cluster-randomised trial in antenatal clinics (ANC) in Kisumu County and Homa Bay County, western Kenya. The trial ran from November 11, 2019, to December 10, 2020, in 15 health facilities (originally 18, but three were excluded due to the COVID-19 pandemic). The study design, eligibility criteria, sample size, and randomisation procedures are described in detail by Barsosio et al [18].

Briefly, the three arms comprised: (1) IPTp with SP (IPTp-SP), (2) IPTp with DP (IPTp-DP), and (3) IPTp-DP plus provider-targeted education (DP-plus). The trial had a 10-month implementation phase and an end-line evaluation that included ANC exit interviews and home visit follow-ups in the final four months (i.e., from September 8 to December 10, 2020). The household cost analysis was conducted using data from the exit interviews with 1,189 pregnant women (377 in the IPTp-SP group, 408 in the IPTp-DP group, and 404 in the DP-plus group).

### Data collection

Exit interviews were conducted with pregnant women in their second or third trimester using a questionnaire administered face-to-face by trained research assistants, approximately 5-6 months after the start of IPTp implementation. Information about whether the IPTp drugs were given during the ANC visits was obtained by checking the ANC cards during the interviews. The questionnaire was translated into Kiswahili and Dholuo, back-translated to English for verification, pre-tested, and administered using electronic devices.

The questionnaire collected information on demographics, sources of healthcare, care-seeking behaviour, health services received, and payments made during the current visit. Participants were asked about the time they spent travelling and waiting at the facility, whether an adult accompanied them, and the types of services they received, as well as any out-of-pocket expenses they incurred. Malaria-specific questions focused on whether they received outpatient or inpatient treatment for malaria during the current pregnancy. A positive response was followed by detailed questions about the direct and indirect costs associated with the last episode of outpatient and inpatient care. Direct medical costs included fees paid for registration and consultation, laboratory tests, medications, and other expenses incurred during outpatient visits and hospital admissions. Direct non-medical costs included the fare paid to travel to the health facilities. We also inquired about indirect costs associated with lost productivity by asking about the number of days for which patients and any accompanying person were unable to engage in economic activities while seeking care or during illness. In addition, we inquired about overall healthcare costs in case the participant could not recall the detailed cost items from the last episode of malaria.

The final part of the questionnaire covered ownership of assets and household characteristics based on the Demographic and Health Survey, such as floor materials, number of rooms, type of toilets, cooking fuel, sources of drinking water, and average household income.

### Data analysis

All costs were collected in Kenyan shillings, inflated to 2024 using the GDP deflator and finally converted to US dollars (USD) using an exchange rate of 1 USD=134.7 KSh. Participants were from Kisumu City and Homa Bay County, hence the minimum wages of 13,572.9 KSh (∼101 USD) and 7,240.95 KSh (∼54 USD) per month for a general labourer were used to value the loss in productivity, respectively [19]. We assumed a full working day was lost for each day of care-seeking. Some participants had several episodes of malaria; hence, we only collected the costs for the last episode to minimise recall bias. However, in estimating the total cost per pregnancy, we assumed the treatment costs were the same for each episode. The questionnaire asked participants to list itemised costs, which were summed to obtain the total costs incurred. Additionally, we inquired about the total lump sum cost paid as an option for those who may have forgotten the itemised costs. During the computation of treatment costs, we used the highest of the itemised total and the lump sum.

We used ownership of assets to construct socioeconomic status indices using a principal component analysis. The degree of inequality in the distribution of costs across socioeconomic groups was measured by the concentration index, which ranges between -1 and 1, with a positive (negative) value indicating a higher incidence among the richest (poorest) quintile [20]. The further the value of the concentration index is from zero, the higher the degree of inequality. Inequality was assessed using the “conindex” command in Stata [21]. Catastrophic health expenditure (CHE) was defined as household costs associated with care-seeking for MiP that exceed 10% of household income [22].

Cost data are often skewed; therefore, we used a mixed-effects generalised linear model with a gamma distribution and log-link function to examine factors associated with household costs. We adjusted for trial arm and clustering, and all tests used a two-sided α of 0.05. Analyses were conducted in Stata version 16.1.

### Ethical considerations

The study received ethical clearance from the Liverpool School of Tropical Medicine [18-073] (sponsor), the Kenya Medical Research Institute [KEMRI/SERU/ CGHR/005/3751], Regional Committees for Medical Research Ethics Southeast Norway-REC [2018-2112], and London School of Hygiene and Tropical Medicine in UK-LSHTM REC [17179]. Written informed consent was obtained from all participants before the exit interviews, which were conducted in private to ensure confidentiality. This trial is registered on ClinicalTrials.gov (NCT04160026) and is complete.

## Results

### Description of the study participant

Of the 1,189 pregnant women who participated in the exit interviews (377 in the IPTp-SP, 408 in the IPTp-DP, and 404 in the IPTp-DP plus), sociodemographic characteristics were similar across groups (Table 1). The mean age was 24 years; approximately one-third were on their first pregnancy, and nearly 90% were Luo, the dominant ethnic group in the area. Approximately three-quarters were married or cohabiting; 40% and 30% had completed primary and secondary education, respectively. Women in the two IPTp-DP arms were relatively poorer than those in the IPTp-SP arm based on ownership of assets and household characteristics.

**Table 1.** Characteristics of pregnant women interviewed.

|  | All | By trial arms |  |  |
| --- | --- | --- | --- | --- |
| Sociodemographic characteristics | N=1,189 | SP (N=377) | DP (N=408) | DP Plus (N=404) |
| Average age in years (sd) | 24 (5.4) | 25 (5.4) | 24 (5.2) | 25 (5.6) |
| **Marital status (%)** |  |  |  |  |
| Single | 265 (22%) | 88 (23%) | 90 (22%) | 87 (22%) |
| Married/cohabiting | 908 (76%) | 286 (76%) | 313 (77%) | 309 (76%) |
| Divorced/widowed | 16 (1%) | 3 (1%) | 5 (1%) | 8 (2%) |
| **Education level attained (%)** |  |  |  |  |
| None | 209 (18%) | 62 (16%) | 72 (18%) | 75 (19%) |
| Primary | 477 (40%) | 147 (39%) | 159 (39%) | 171 (42%) |
| Secondary | 353 (30%) | 107 (28%) | 133 (33%) | 113 (28%) |
| Post-secondary | 150 (12%) | 61 (16%) | 44 (11%) | 45 (11%) |
| **Ethnicity (%)** |  |  |  |  |
| Luo | 1040 (88%) | 331 (88%) | 359 (88%) | 350 (87%) |
| Luhya | 68 (6%) | 22 (6%) | 27 (7%) | 19 (5%) |
| Kisii | 32 (3%) | 12 (3.2%) | 12 (3%) | 8 (2%) |
| Others | 49 (4%) | 12 (3%) | 10 (2%) | 27 (7%) |
| **Religion (%)** |  |  |  |  |
| Christians | 1173 (99%) | 369 (98%) | 405 (99%) | 399 (99%) |
| Other | 16 (1%) | 8 (2%) | 3 (1%) | 5 (1%) |
| **Gravidity** |  |  |  |  |
| Primigravidae | 397 (33%) | 118 (31%) | 143 (35%) | 136 (34%) |
| **Socioeconomic status (%)** |  |  |  |  |
| Poorest | 238 (20%) | 57 (15.1%) | 94 (23.0%) | 87 (21.5%) |
| 2<sup>nd</sup> Poorest | 238 (20%) | 47 (12.5%) | 83 (20.3%) | 108 (26.7%) |
| Middle | 238 (20%) | 80 (21.2%) | 78 (19.1%) | 80 (19.8%) |
| 2<sup>nd</sup> Least poor | 238 (20%) | 93 (24.7%) | 75 (18.4%) | 70 (17.3%) |
| Least poor | 237 (20%) | 100 (26.5%) | 78 (19.1%) | 59 (14.6%) |
| **Reasons for clinic visit (%)** |  |  |  |  |
| Routine ANC visit | 1096 (92%) | 360 (95%) | 367 (90%) | 369 (91%) |
| Registration for ANC | 84 (7%) | 11 (3%) | 40 (10%) | 33 (8%) |
| Other | 9 (1%) | 6 (2%) | 1 (0%) | 2 (1%) |
| **Trimester of pregnancy** |  |  |  |  |
| 3<sup>rd</sup> Trimester | 1135/1178 (96%) | 369/376 (98%) | 375/400 (92%) | 391/402 (97%) |
| **# of other facilities visited for ANC** |  |  |  |  |
| 1 | 300 (25%) | 124 (33%) | 98 (24%) | 78 (19%) |
| 2 or more | 66 (6%) | 22 (6%) | 35 (9%) | 10 (2%) |
| Use of SP during the current pregnancy |  | 368 (98%) | 8 (2%) | 17 (4%) |
| Current visit for ANC but not given IPTp drugs | 72/1096 (7%) | 21/360 (5%) | 29/367 (8%) | 22/369 (7%) |
| **ANC visits recorded in the card (%)** |  |  |  |  |
| 1 visit | 142 (12%) | 26 (7%) | 63 (15%) | 53 (13%) |
| 2 visits | 250 (21%) | 78 (21%) | 99 (24%) | 73 (18%) |
| 3 visits | 321 (27%) | 99 (26%) | 125 (31%) | 97 (24%) |
| 4 visits | 215 (18%) | 76 (20%) | 71 (17%) | 68 (17%) |
| 5 visits | 147 (12%) | 54 (14%) | 33 (8%) | 60 (15%) |
| More than 5 visits | 113 (10%) | 45 (11%) | 16 (4%) | 53 (13%) |

SP: sulfadoxine-pyrimethamine, DP: dihydroartemisinin-piperaquine, DP-plus: dihydroartemisinin-piperaquine plus provider education, ANC: antenatal care

Ninety-six percent of women were in their third trimester. Antenatal cards showed that 7% of women in the IPTp-SP arm and twice as many in the IPTp-DP and DP-plus arms were attending routine ANC for the first time at the time of the exit interview. Approximately one-third of the participants had visited other health facilities for ANC services; this proportion was higher in the IPTp-SP arm. Two women assigned to the IPTp-SP arm also used IPTp-DP. Most women attended facilities for routine ANC visits on the interview day; however, 9.8% in the IPTp-DP and 8.2% in the IPTp-DP plus arms visited health facilities to register for ANC (Table 1).

### History of care-seeking for malaria

Of the 1,189 participants, 247 (21%) self-reported having received treatment for malaria during the current pregnancy. Of these, 212 (86%) were treated as outpatients (209 in the last visit), 24 (10%) as inpatients, and 11 (4%) did not specify the type of care received (Fig 1). By trial arm, malaria care-seeking occurred in 99 women (26%) in the IPTp-SP arm, 85 (21%) in the IPTp-DP arm, and 63 (16%) in the IPTp-DP plus arm (Table 2 and Fig 1). About 58% of outpatient malaria care was sought from study clinics, 27% from other health facilities, and 10% from private clinics. About 60% of the 24 participants who reported inpatient care were admitted to public hospitals.

**Table 2.** Care-seeking for malaria.

|  | All | By trial arms |  |  |
| --- | --- | --- | --- | --- |
| Malaria-related care | N=1189 | SP (N=377) | DP (N=408) | DP Plus (N=404) |
| Number of women treated for malaria | 247 (21%) | 99 (26%) | 85 (21%) | 63 (16%) |
| Number of women treated for malaria during the last OPD visit | 209 (18%) | 84 (22%) | 73 (18%) | 52 (13%) |
| Treated for 1 episode | 158/209 (75%) | 63/84 (75%) | 53/73 (73%) | 42/52 (81%) |
| Treated for 2 or more episodes | 51/209 (25%) | 21/84 (25%) | 20/73 (27%) | 10/52 (19%) |
| Number of women who sought inpatient care for malaria | 24 (2%) | 12 (3%) | 7 (2%) | 5 (1%) |
| **Source of OPD malaria care** |  |  |  |  |
| The study facility | 131 (58%) | 51 (57%) | 41 (53%) | 39 (68%) |
| Another facility | 61 (27%) | 27 (30%) | 23 (30%) | 11 (19%) |
| Private clinic | 22 (10%) | 9 (10%) | 11 (14%) | 2 (3%) |
| Pharmacy | 8 (4%) | 3 (3%) | 2 (3%) | 3 (5%) |
| Accompanied by an adult person for OPD | 91/224 (41%) | 39/90 (43%) | 37/77 (40%) | 15/57 (26%) |
| Average # of days unable to work (OPD) | 4.5 (4.3) | 4.2 (3.6) | 5.6 (4.8) | 3.5 (4.4) |
| **Source of inpatient malaria care** |  |  |  |  |
| Public hospital | 14/24 (58%) | 7/12 (58%) | 3/7 (43%) | 4/5 (80%) |
| Private hospital | 10/24 (42%) | 5/12 (42%) | 4/7 (57%) | 1/5 (20%) |
| Average admission days (sd) | 3.3 (2.2) | 2.7 (1.5) | 3.7 (1.9) | 4 (4) |
| Accompanied by an adult person for IPD | 19/24 (79%) | 10/12 (83%) | 4/7 (57%) | 5/5 (100%) |
| Average # of days unable to work (IPD) | 9.2 (5) | 8 (5) | 11 (5.5) | 10 (4.4) |

SP: sulfadoxine-pyrimethamine, DP: dihydroartemisinin-piperaquine, DP-plus: dihydroartemisinin-piperaquine plus provider education, ANC-antenatal care, OPD-Outpatient department, IPD-Inpatient department, sd-standard deviation,

**Fig 1.**
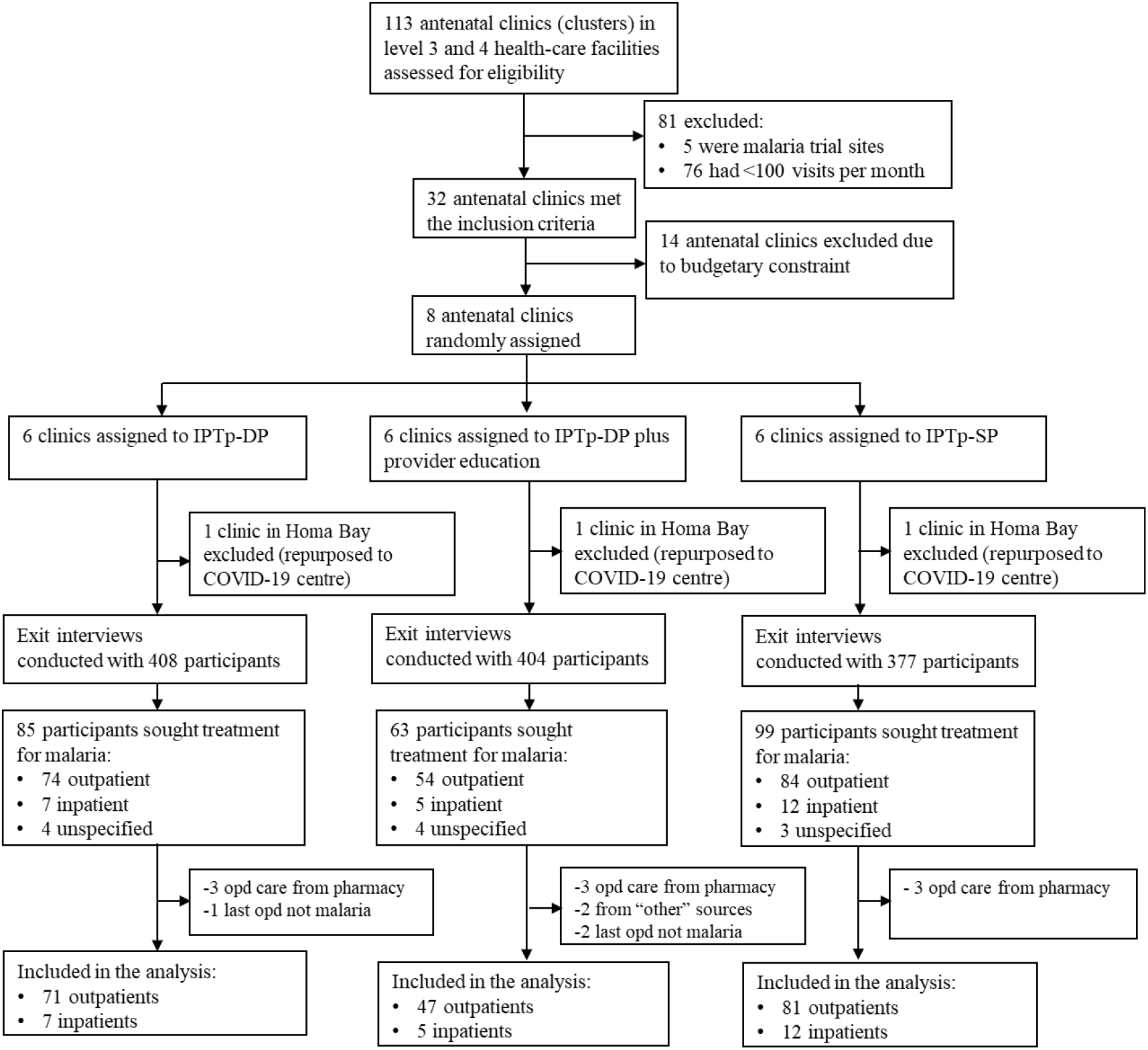
Trial profile. OPD-outpatient department, SP: sulfadoxine-pyrimethamine, DP: dihydroartemisinin-piperaquine, DP-plus: dihydroartemisinin-piperaquine plus provider education

### Household costs associated with MiP

Of the 212 outpatient episodes, 13 were excluded: three to restrict analysis to the last outpatient visit to malaria (minimising recall bias) and 10 who sought care from pharmacies, traditional healers, or relatives (to maintain comparability). The economic analysis, therefore, included 199 outpatient and 24 inpatient episodes (Fig 1).

The overall mean outpatient cost for treating one episode of MiP, regardless of IPTp strategy used, was USD 24.9; the median was USD 20.3. The mean inpatient MiP cost was USD 57.1; the median was USD 59.6. Productivity losses from waiting time and incapacitation contributed the largest share of costs (Table 3), accounting for about 87% of outpatient and 70% of inpatient costs, respectively. Mean outpatient costs were equivalent to 6 days’ minimum wages for a general labourer in Kisumu and 12 days’ wages in Homa Bay. Mean inpatient costs were equivalent to half a month’s minimum wages in Kisumu and one month’s wages in Homa Bay.

**Table 3.** Mean and median costs (USD) for malaria treatment in pregnancy.

| **Outpatient care** |  |  |  |
| --- | --- | --- | --- |
| **Cost category** | **N** | **Mean (sd)** | **Median (IQR)** |
| Registration | 19 | 0.7 (0.6) | 0.5 (0.3-1.0) |
| Consultation | 9 | 4.0 (4.4) | 2.7 (0.9-3.6) |
| Laboratory test | 30 | 1.6 (1.0) | 1.3 (0.9- 2.4) |
| Drugs | 29 | 2.6 (2.1) | 2.3 (0.9-3.6) |
| Other costs | 5 | 2.3 (1.6) | 2.3 (1.4-3.2) |
| Food, drinks etc | 38 | 5.1 (6.1) | 2.9 (1.8-6.3) |
| Transport | 80 | 0.9 (0.7) | 0.5 (0.5-0.9) |
| Indirect cost | 199 | 21.7 (16.7) | 16.3 (8.7-32.6) |
| Mean total |  | 24.9 (18.7) | 20.3 (12.2-35.3) |
| **Inpatient care** |  |  |  |
| Registration | 4 | 1.6 (1.3) | 1.8 (0.5-2.7) |
| Hospitalisation | 2 | 3.1 (1.9) | 3.2 (1.8-4.5) |
| Laboratory test | 4 | 3.7 (3.6) | 2.5 (1.4-6.1) |
| Drugs | 2 | 20.5 (2.9) | 20.5 (18.4-22.5) |
| Food, drinks | 14 | 9.3 (10.5) | 4.3 (1.8-18.0) |
| Other costs | 2 | 7.1 (1.4) | 7.1 (6.1-8.1) |
| Transport | 20 | 1.6 (2.0) | 0.9 (0.5-1.6) |
| Indirect cost | 24 | 39.8 (19.2) | 36.6 (33.7-61.1) |
| **Mean total** |  | **57.1 (33.4)** | **59.6 (35.9-70.8)** |

SP: sulfadoxine-pyrimethamine, DP: dihydroartemisinin-piperaquine, DP-plus: dihydroartemisinin-piperaquine plus provider education, sd-standard deviation, IQR-interquartile range

**Table 4.** Mean and median cost estimates (USD) for outpatient and inpatient care for malaria.

| **Outpatient care** |  |  |  |  |  |  |  |  |  |
| --- | --- | --- | --- | --- | --- | --- | --- | --- | --- |
|  | **SP** |  |  | **DP** |  |  | **DP Plus** |  |  |
| **Cost category** | **N** | **Mean (sd)** | **Median (IQR)** | **N** | **Mean (sd)** | **Median (IQR)** | **N** | **Mean (sd)** | **Median (IQR)** |
| Registration | 6 | 0.4 (0.1) | 0.5 (0.3-0.5) | 8 | 0.8 (0.7) | 0.5 (0.3-1.1) | 5 | 0.8 (0.6) | 0.9 (0.6-0.9) |
| Consultation | 3 | 5.7 (6.9) | 3.2 (0.5-13.5) | 5 | 3.2 (3.3) | 2.7 (0.9-2.7) | 1 | 3.6 | 3.6 |
| Laboratory test | 9 | 1.8 (1.4) | 0.9 (0.9- 3.2) | 13 | 1.6 (1.0) | 1.4 (0.9-2.7) | 8 | 1.3 (0.5) | 1.4 (0.9-1.8) |
| Drugs | 13 | 2.6 (2.2) | 2.3 (0.9-4.1) | 12 | 2.5 (1.8) | 2.3 (1.3-3.4) | 4 | 3.4 (2.5) | 2.9 (1.6-5.2) |
| Other costs | 1 | 3.1 | 3.2 | 3 | 2.4 (2.0) | 2.3 (0.5-4.5) | 1 | 1.4 | 1.4 |
| Food and drinks | 19 | 4.3 (3.9) | 3.6 (0.9-7.2) | 14 | 6.8 (8.8) | 2.9 (1.8-6.3) | 5 | 3.5 (3.3) | 2.3 (1.8-4.1) |
| Transport | 29 | 0.6 (0.4) | 0.5 (0.5-0.9) | 36 | 1.1 (0.8) | 0.9 (0.5-1.8) | 15 | 0.9 (0.7) | 0.9 (0.5-0.9) |
| Indirect cost | 81 | 22.3 (15.9) | 17.4 (12.2-32.6) | 71 | 23.2 (16.8) | 19.5 (10.9-32.6) | 47 | 18.4 (17.6) | 12.2 (4.1-32.6) |
| **Total** |  | **25.5 (16.7)** | **20.4 (14.5-34.4)** |  | **27.3 (20.0)** | **20.5 (12.2-36.6)** |  | **20.3 (19.4)** | **13.1 (4.1-32.6)** |
| **Inpatient care** |  |  |  |  |  |  |  |  |  |
| **Cost category** | **N** | **Mean** | **Median (IQR)** | **N** | **Mean** | **Median (IQR)** | **N** | **Mean** | **Median (IQR)** |
| Registration | 2 | 0.5 (0.5) | 0.5 (0.2-0.9) | 2 | 2.7 | 2.7 | 0 | - | - |
| Consultation | 1 | 4.5 | 4.5 | 1 | 1.8 | 1.8 | 0 | - | - |
| Laboratory test | 2 | 2.0 (1.6) | 2.0 (0.9- 3.1) | 1 | 1.8 | 1.8 | 1 | 9.0 | 9.0 |
| Drugs | 1 | 18.4 | 18.4 | 1 | 22.5 | 22.5 | 0 | - | - |
| Other costs | 1 | 6.1 | 6.1 | 1 | 8.1 | 8.1 | 0 | - | - |
| Transport | 11 | 1.1 (0.8) | 0.9 (0.4-0.9) | 5 | 2.6 (3.6) | 0.9 (0.9-1.8) | 4 | 1.7 (1.9) | 1.0 (0.5-2.9) |
| Indirect cost | 12 | 34.7 (17.2) | 36.6 (25.5-36.6) | 7 | 42.2 (23.1) | 34.7 (32.6-65.1) | 5 | 48.8(18.1) | 52.9 (36.6-65.1) |
| **Total** |  | **45.6 (17.3)** | **41.8 (35.1-64.1)** |  | **77.1 (52.0)** | **79.7 (34.7-94.8)** |  | **56.9 (20.7)** | **66.4 (41.9-68.3)** |

SP: sulfadoxine-pyrimethamine, DP: dihydroartemisinin-piperaquine, DP-plus: dihydroartemisinin-piperaquine plus provider education, ANC: antenatal care, sd- standard deviation, IQR-Interquartile range

The mean outpatient costs for MiP among women receiving IPTp-SP, IPTp-DP, and IPTp-DP plus were USD 25.5, USD 27.3, and USD 20.3, respectively. As cost distributions were slightly right-skewed, the corresponding median outpatient costs were USD 20.4 for IPTp-SP, USD 20.5 for IPTp-DP, and USD 13.1 for IPTp-DP plus (Table ). In the adjusted model, each year increase in maternal age was associated with a 5% increase in outpatient costs (β=0.045, p=0.001), and being accompanied by an adult increased costs by 43% (β=0.425, p=0.005).

Of the 24 pregnant women who reported to have been admitted to hospital due to MiP, the mean costs were USD 45.6 (median USD 41.8) for IPTp-SP, USD 77.1 (median USD 79.7) for IPTp-DP, and USD

56.9 (median USD 66.4) for IPTp-DP plus (Table ). More than half of the patients in the IPTp-DP arm were admitted to private hospitals, which could explain the higher costs.

### Equity in the distribution of costs and catastrophic health expenditure

We found no differences in the distribution of outpatient costs by socioeconomic status. For the entire cohort, the concentration index was 0.02 (p=0.571). By treatment arm, concentration indices were 0.06 (p=0.172) for IPTp-DP plus, -0.003 (p=0.969) for IPTp-DP, and 0.04 (p=0.427) for IPTp-SP (Fig 2).

**Fig 2.**
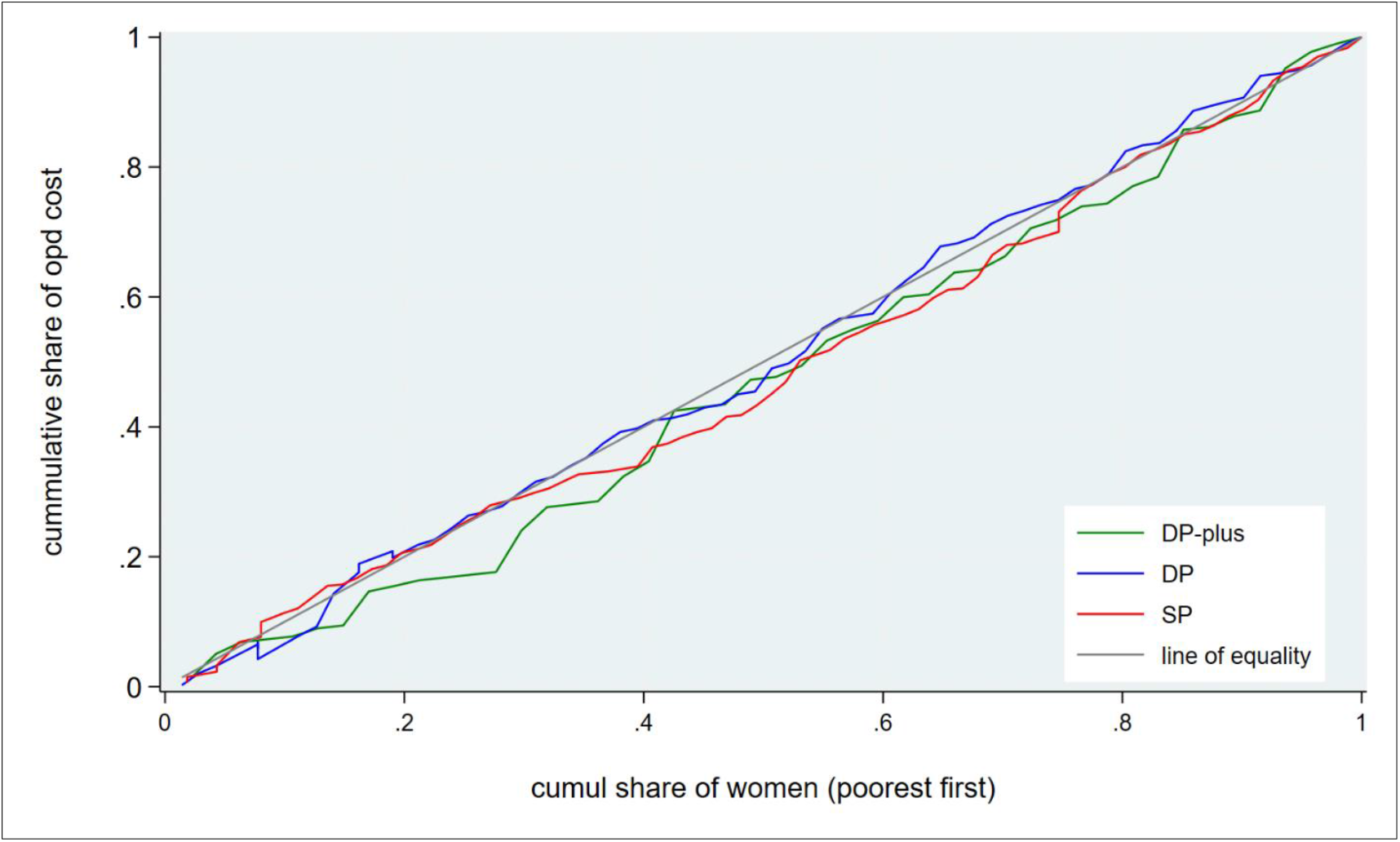
Concentration curve for the distribution of outpatient malaria costs. SP-sulfadoxine-pyrimethamine, DP-dihydroartemisinin-piperaquine, DP-plus- dihydroartemisinin-piperaquine combined with provider education

Only 111 participants with outpatient episodes had income information; 8% experienced catastrophic health expenditure (CHE). The degree of inequality in the distribution of CHE was very small and not statistically significant (concentration index 0.01, p=0.304). However, there were indications that CHE occurred predominantly in the middle and second poorest quintiles, indicating a relatively higher economic burden among poorer households (Fig 3). CHE distribution by treatment arm was not analysed owing to small sample sizes. For inpatient care, among 12 participants with income information, 11 (92%) experienced CHE, which was concentrated in the poorest (46%), second poorest (9%), and middle (18%) quintiles.

**Fig 3.**
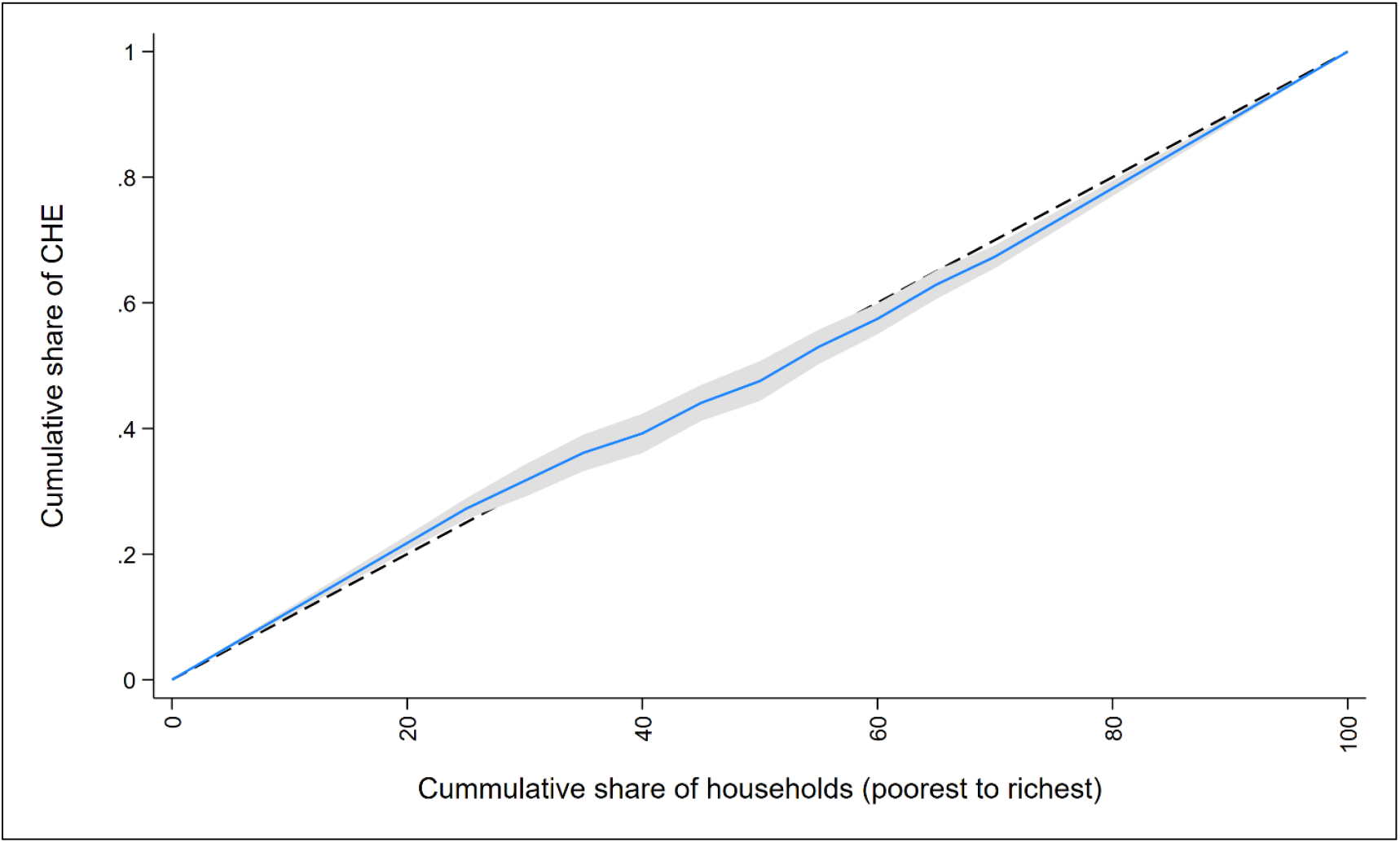
Concentration curve for the distribution of CHE (catastrophic health expenditure) Dotted line represents the line of equality, and the grey area is the 95% confidence interval

## Discussion

In this study examining the household economic burden of malaria in pregnancy among HIV-negative pregnant women in western Kenya, we found that the mean household costs per episode were substantial: estimated at USD 24.9 for outpatient care and USD 57.1 for inpatient care. These costs represented six days of minimum wages for a general labourer in Kisumu and 12 days in Homa Bay for outpatient care, while inpatient costs equated to half a month’s wages in Kisumu and a full month’s wages in Homa Bay [19]. Average indirect costs, comprising productivity losses from travel time, waiting, incapacitation, and accompaniment time, accounted for 87% and 70% of average total outpatient and inpatient costs, respectively.

Our secondary findings revealed that approximately one in five pregnant women (20.8%) self-reported seeking care for one or more episodes of malaria during the study period. The incidence of catastrophic health expenditure (CHE) was 8% for outpatient care overall, with CHE concentrated among middle and second-poorest wealth quintiles. For inpatient care, 92% of households with income data experienced CHE, with three-quarters belonging to the middle, second-poorest, and poorest quintiles. While mean outpatient costs did not differ significantly by socioeconomic status (concentration index 0.02, p=0.571), the burden relative to household resources disproportionately affected poorer households.

Our cost estimates align with recent findings from sub-Saharan Africa. Cirera and colleagues [11] reported mean outpatient MiP costs (adjusted to 2024) per episode of USD 25.8 in Democratic Republic of Congo, USD 18.9 in Madagascar, USD 34.6 in Mozambique, and USD 21.4 in Nigeria, with indirect costs representing 73-97% of total costs. Their inpatient estimates (USD 40.5-69.6) were similarly comparable to our findings. Evidence from outside Africa is limited to Colombia, where, although malaria treatment is provided free of charge, pregnant women with malaria incurred median costs of USD 16.3 per outpatient visit and USD 26.7 per admission (2011 prices); transportation and productivity losses accounted for most of these costs [23]. The high proportion of indirect costs in our study reflects the substantial productivity losses experienced by pregnant women and their families when seeking care for malaria. Another study in Nigeria in 2020, estimated average direct medical and non-medical MiP costs at USD 14.9 and USD 23.8 for outpatient care [24]. Earlier studies from West Africa reported lower costs: Duval et al (2022) [14] estimated outpatient costs at USD 5.0 in Burkina Faso and USD 1.0 in The Gambia, with indirect costs of USD 15.1 and USD 5.2, respectively. In Mozambique, an earlier study estimated the outpatient MiP cost at USD 8.4, of which indirect costs accounted for 72%, while inpatient costs were USD 47.8, with indirect costs contributing 25% [25]. These variations likely reflect differences in wage levels, treatment practices, distances from health facilities, costing perspectives, and the time elapsed since these studies were conducted, with minimum monthly wages in The Gambia (USD 25) and Burkina Faso (USD 51) being considerably lower than in our Kenyan study sites (USD 54-101) [26, 27].

Household costs accrue with each episode of malaria; effective prevention of malaria in pregnancy therefore also reduces the economic burden on households. The comparative cost-effectiveness of the IPTp strategies evaluated in this trial is reported separately.

Our study had several limitations. First, malaria episodes were self-reported and could not be verified through parasitological confirmation, which may have led to misclassification. Second, recall bias may have affected the cost estimates, as women reported expenses incurred up to several months before the interview. Third, cost data were collected only for the most recent outpatient MiP episode; therefore, identical treatment costs were assumed for participants with multiple episodes, which may have led to misestimation of the true economic burden. Fourth, we assumed that full productive days were lost during care-seeking, which may have overestimated indirect costs. Conversely, costs may have been underestimated if women lost additional productive time due to delayed care-seeking. Cirera et al (2023) reported waiting times of 1-2 hours and transport times of 1-3 hours in their multi-country study [11]. Finally, we lacked comprehensive household consumption data, necessitating the use of income data for CHE calculations. Income-based CHE estimates are less reliable in rural settings where informal sector work and home production are common [20, 28] and income underreporting may have further limited these estimates [29].

### Policy implications

Despite Kenya’s policy of free reproductive and child health services, pregnant women incurred costs for registration, drugs, consultations, and laboratory tests for malaria [30]. Our results indicate that implementation gaps persist, with households bearing significant out-of-pocket costs for malaria treatment during pregnancy. Strengthening social protection mechanisms and ensuring effective implementation of user fee exemptions for pregnant women are essential to reduce the economic burden of malaria in pregnancy.

## Conclusion

Household costs associated with malaria care-seeking during pregnancy in western Kenya imposed a substantial economic burden relative to local wages, with 8% of households experiencing catastrophic health expenditure for outpatient care and all but one woman (with income data) who sought inpatient care. While mean costs did not vary by socioeconomic status, the burden relative to household resources was greater among poorer households. These findings support effective prevention of malaria in pregnancy and strengthened implementation of free maternal health services.

## Data Availability

Data dictionaries and de-identified participant data can be shared by ATM on request. However, such requests must be reviewed by the Kenya Medical Research Institute Scientific Ethics and Review Unit and must conform to the Kenya Data Protection Act of 2019.

## Article Information

## Acknowledgments

We thank the women who participated in the study and the dedicated study staff.

## Contributors

ATM-data curation, formal analysis and writing original draft. SF, KH, JH, FOtK and BR contributed to the statistical analysis and validation and supported ATM with manuscript revisions, editing and data interpretation. JH, JW, SF, FOtK and BR conceived the study. JH, JW, HCB, SK, SF and BR were responsible for the final study design. SF, KH, HCB, FO, AK, MAO, DW, BO, EO, MT, SK, MM, JL, FM, DW, DM, VM, KP, KM, JW, JH, FOtK and BR contributed to funding and data acquisition. HCB, JH, SF and FOtK directly accessed and verified the underlying data reported in this article. All authors contributed to critical revisions of the manuscript. All authors had access to all the data in the study and accept responsibility for the decision to submit for publication.

